# Assessing Compliance with Smoke-Free Laws in Purbakhola Rural Municipality, Nepal: A Cross-Sectional Observational Study

**DOI:** 10.1101/2025.11.05.25339641

**Authors:** Bhakta Bahadur KC, Ananda Bahadur Chand, Amit Bam, Tara Singh Bam

**Author notes:** Corresponding Author: Bhakta Bahadur KC.

## Abstract

**Introduction:** Despite Nepal’s comprehensive tobacco control legislation, evidence on its implementation at the local level, particularly in rural areas, remains scarce. This study assessed compliance with smoke-free laws in public places within Purbakhola Rural Municipality, Palpa, Nepal.

**Methods:** A cross-sectional observational survey was conducted from May, 2025, across all six wards of the municipality. A total of 219 public places across 14 categories were selected. Compliance was measured using an adapted observational checklist assessing six key indicators: presence of signage, active smoking, astryas, designated smoking areas, and cigarette butt litter. Data were analyzed using IBM SPSS 31.

**Results:** Compliance was high in institutional settings such as health facilities (100.0% on most indoor measures), educational institutions (100.0%), and government offices (88.8-100.0%). Significant gaps were identified in the hospitality sector, where only 45.0% of restaurants were free from active indoor smoking and only 37.5% had no visible ashtrays. A critical finding was the systemic lack of compliant no-smoking signage, even in high-compliance settings, with 54.5% of health facilities non-compliant indoors and 100.0% of government offices non-compliant outdoors. The visible ashtrays were the strongest predictor of smoking, with indoor ashtrays associated with dramatically higher odds of observed smoking. In contrast, no statistically significant association was found for signage, though a promising signal showed 0% smoking in venues with indoor signage present.

**Conclusion:** This study reveals a significant implementation gap of tobacco control in a rural municipality of Nepal The findings underscore the urgent need for a dual strategy: rigorous enforcement to remove ashtrays—a major environmental cue for smoking—and a concerted campaign to ensure the universal installation of standardized ‘No Smoking’ signage to align practice with policy and protect public health.

## INTRODUCTION

Tobacco use remains one of the world’s most significant public health threats, responsible for over 8 million deaths annually(1, 2). The burden of tobacco-related disease is disproportionately borne by low- and middle-income countries like Nepal, where tobacco smoking prevalence is 46.3% among men and 11.6% among women, contributing to over 39,000 deaths yearly(3–7). Secondhand smoke poses serious health risks to non-smokers, including increased risks of heart disease, lung cancer, and respiratory illnesses. Among of the 8 million global death from tobacco use, 1.6 million deaths are non-smokers exposed to second-hand smoke (SHS)(1, 2).

In response, Nepal ratified the World Health Organization Framework Convention on Tobacco Control (WHO FCTC) in 2006(8). The WHO FCTC Article 8 and its guidelines establish 100% smoke-free environments as the best practice for protecting citizens from SHS(9). Similarly, the MPOWER, the WHO’s technical assistance package of evidence-based policies, also identifies the adoption of 100% smoke-free policies as a critical strategy to reversing the tobacco epidemic. The enactment of the Tobacco Product (Control and Regulatory) Act and Rule in 2011, by Nepal which prohibits smoking in all public places and workplaces(10–12). As per Nepal’s tobacco laws the public places are mainly categorized into 12 entities including indoor and outdoor of government offices, health facilities, schools, restaurants, bars, hotels etc. While the legislation exists, compliance and enforcement of this smoke free public places remain challenges, particularly at the local levels.

In this context, we don’t know the extent of the compliance of smoke free law in local levels. Thus, this study aimed to assess compliance with smoke-free laws within the diverse public places of Purbakhola Rural Municipality, Palpa, Nepal to identify gaps and inform local enforcement and policy strategies.

## METHODS

### Study Design and Setting

A cross-sectional observational study was conducted from May, 2025, in all six wards of Purbakhola Rural Municipality where the population is 16,052). This rural municipality is located in hilly region of Lumbini province of Nepal where the diverse ethnic group live. The study protocol and tools were adapted from the John Hopkins Bloomberg Initiative’s guide for assessing compliance with smoke-free laws(13).

### Operational definitions

#### Compliance

It is the degree to which the Tobacco Product (Control and Regulatory), Act 2011 Nepal is being obeyed.

#### Smoke-free legislation

The law that prohibits smoking in a specified area as per the Tobacco Product (Control and Regulatory), Act 2011 Nepal

#### Public Places

All the places that are defined as public places in the Tobacco Product (Control and Regulatory), Act 2011 Nepal

#### Smoking

Inhaling or exhaling the smoke of tobacco and also includes keeping or controlling any flamed tobacco products.

#### Active smoking

Active smoking in a public place was marked as present if anyone was seen smoking during the researcher’s visit at the public place being observed for the study.

#### ‘No smoking’ signage

According to Tobacco Product (Control and Regulatory), Act 2011 Nepal every public place shall arrange to display a caution notice “Smoking prohibited area, It is a Punishable Offence” in Nepali language at the entrance and in one or more places inside a public place. The size of a caution notice board in a public place shall be at least 40 centimeters * 20 centimeters. Every public place shall arrange to display the caution notice in red letters against a white background or in white letters against a red background with a no-smoking sign.

#### Cigarette buts, bidi ends, or ashes

Cigarette buts, bidi ends, or ashes were marked as present as evidence of previous smoking in a particular public place if any of these was noticed during the researcher’s visit at the public place being observed for the study.

#### Smoking aids

According to Tobacco Product (Control and Regulatory), Act 2011 Nepal, no smoking aids such as ashtrays, ashbins, matchboxes, or lighters can be kept in public places

#### Sampling

A comprehensive list of public places was obtained from the rural municipality office and verified through field visits. Public places were categorized into 14 types (e.g., health facilities, schools, restaurants, government offices, shops, public transport). All 219 identified public places were included. Oral consent was obtained from managers or owners at each venue of public places to observe and take photos.. All collected data were kept confidential and used solely for the purpose of this study.

#### Data Collection and Tool

Trained field enumerators used a standardized checklist to observe each venue for 7-10 minutes during peak business hours. The tool assessed following six compliance indicators:

Presence of no-smoking signage (e.g. written as Smoking Prohibited Zone). Absence of active smoking.

Absence of smoking aids (e.g., ashtrays, lighter, matches). Absence of designated smoking zone.

Absence of cigarette butt litter.

Absence of tobacco product advertisements at point-of-sale.

Photographic evidence was collected to support observations. The supervisor validated data collection by accompanying teams to over 25% of sites as well as by checking data.

#### Data entry and Analysis

Data were entered using a double-entry system to ensure accuracy and analyzed using IBM SPSS version 31. Descriptive statistics were computed to determine compliance rates for each indicator across different categories of public places.

## RESULTS

### Type and Number of Public Places Observed

A total of 219 public places were observed. The sample included health facilities (n=11), government offices (n=9), educational institutions (n=33), restaurants (n=40), shops/malls (n=46), hotels (n=12), and others (n=68)(Table 1)

**Table 1:** Type and Number of Public Places Observed.

| Category of Public Place | Number Surveyed | Percentage of Total |
| --- | --- | --- |
| Shops/Malls | 46 | 21.0% |
| Restaurants / Khaja Ghar | 40 | 18.3% |
| Educational Institutions (total) | 33 | 15.1% |
| Primary/Secondary Schools | 31 | - |
| Colleges/Universities | 2 | - |
| Places of Worship | 27 | 12.3% |
| Others (Furniture shops, Garages) | 12 | 5.5% |
| Hotels/Lodges | 12 | 5.5% |
| Health Facilities | 11 | 5.0% |
| Fitness Centers / Sports Facilities | 11 | 5.0% |
| Private Office Buildings | 10 | 4.6% |
| Government Office Buildings | 9 | 4.1% |
| Public Conveyance (Buses, Jeeps) | 4 | 1.8% |
| Recreation Parks | 2 | 0.9% |
| Bus/Jeep Terminals | 2 | 0.9% |
| **Total** | **219** | **100%** |

### Indoor Compliance with smoke-free laws across different public places

A clear disparity in compliance was evident between formal institutional and hospitality settings. While health facilities, educational institutions, and government offices showed near-perfect adherence to measures prohibiting active smoking, designated smoking areas, and tobacco advertisements, hospitality venues demonstrated marked deficits. The prevalence of active indoor smoking was high in restaurants (55%) and hotels (41.7%), corroborated by the frequent presence of visible ashtrays. The most widespread violation, however, was the absence of compliant signage, which was observed in 54.5% of health facilities and all government offices (Table 2).

**Table 2:**
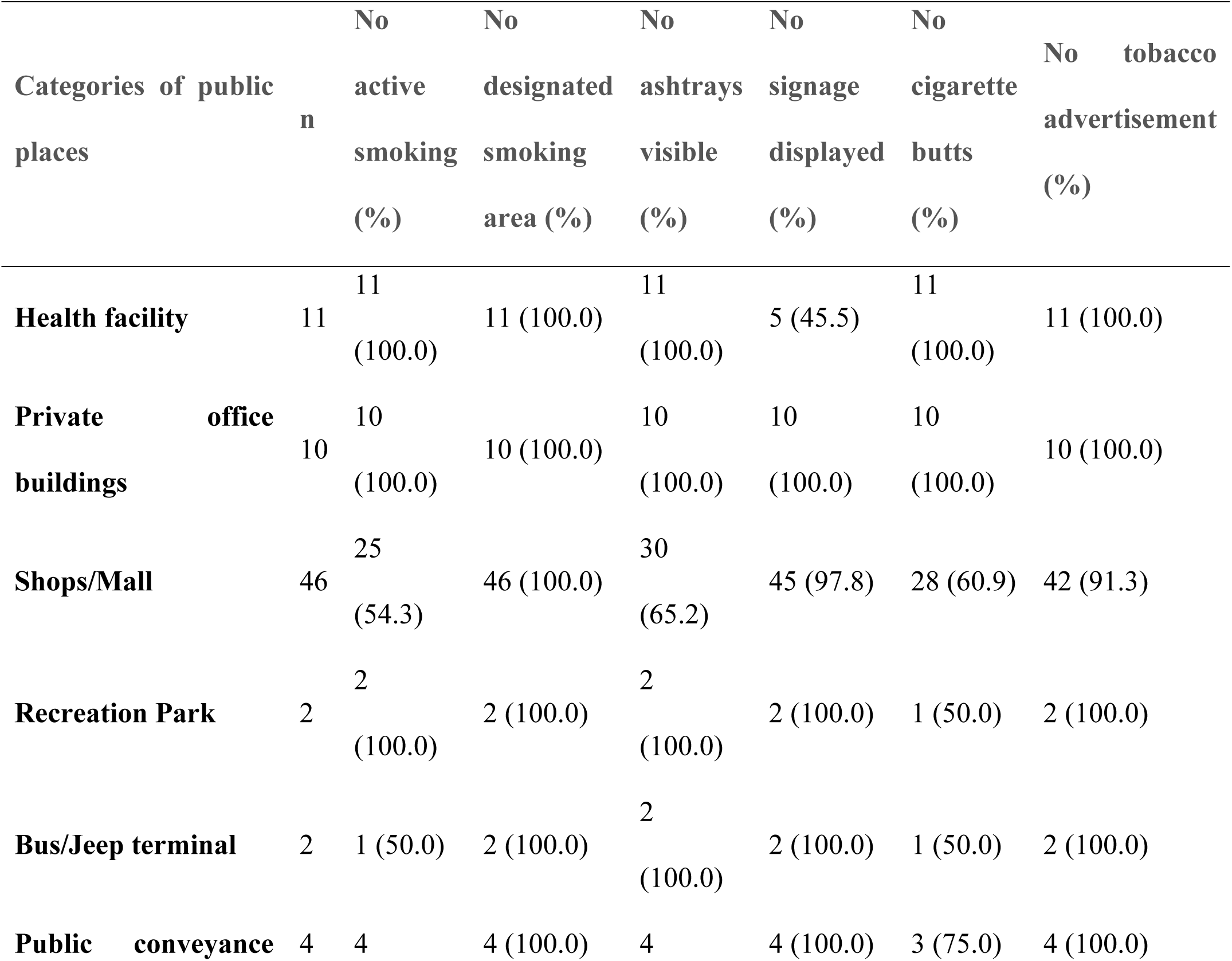
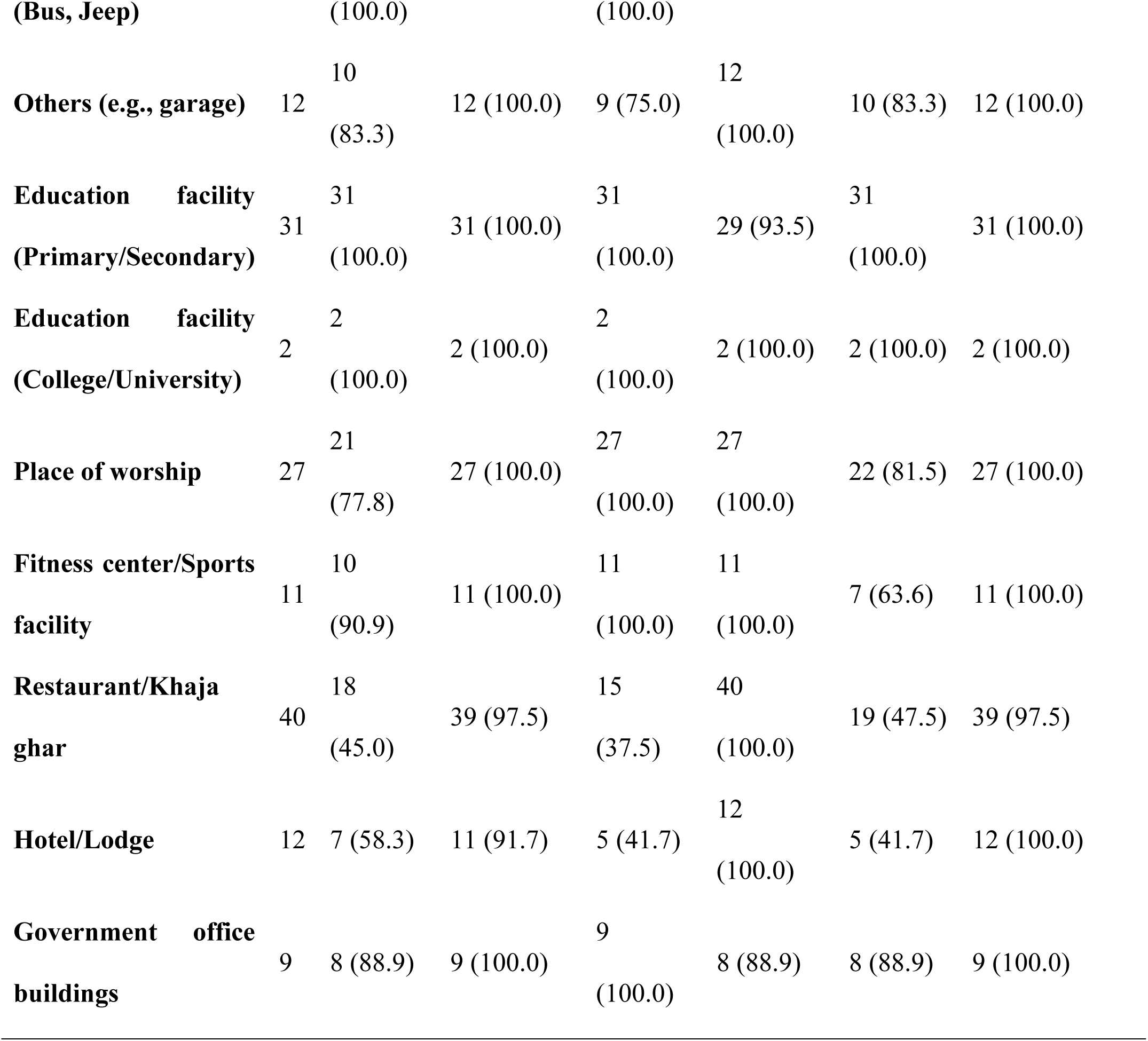
Compliance with smoke-free laws for indoor observations across different public places.

### Outdoor compliance with smoke-free laws across different public places

Outdoor findings within 100 meters of public places revealed similar patterns. A high prevalence of smoking prohibitions was found at health facilities (100.0%) and schools (100%), in stark contrast to restaurants (45.0%). Cigarette butt litter was also a frequent problem, with high non-compliance at government offices (11.1%) and bus terminals (50.0%). Notably, the absence of outdoor signage was nearly universal across all site types (Table 3).

**Table 3:**
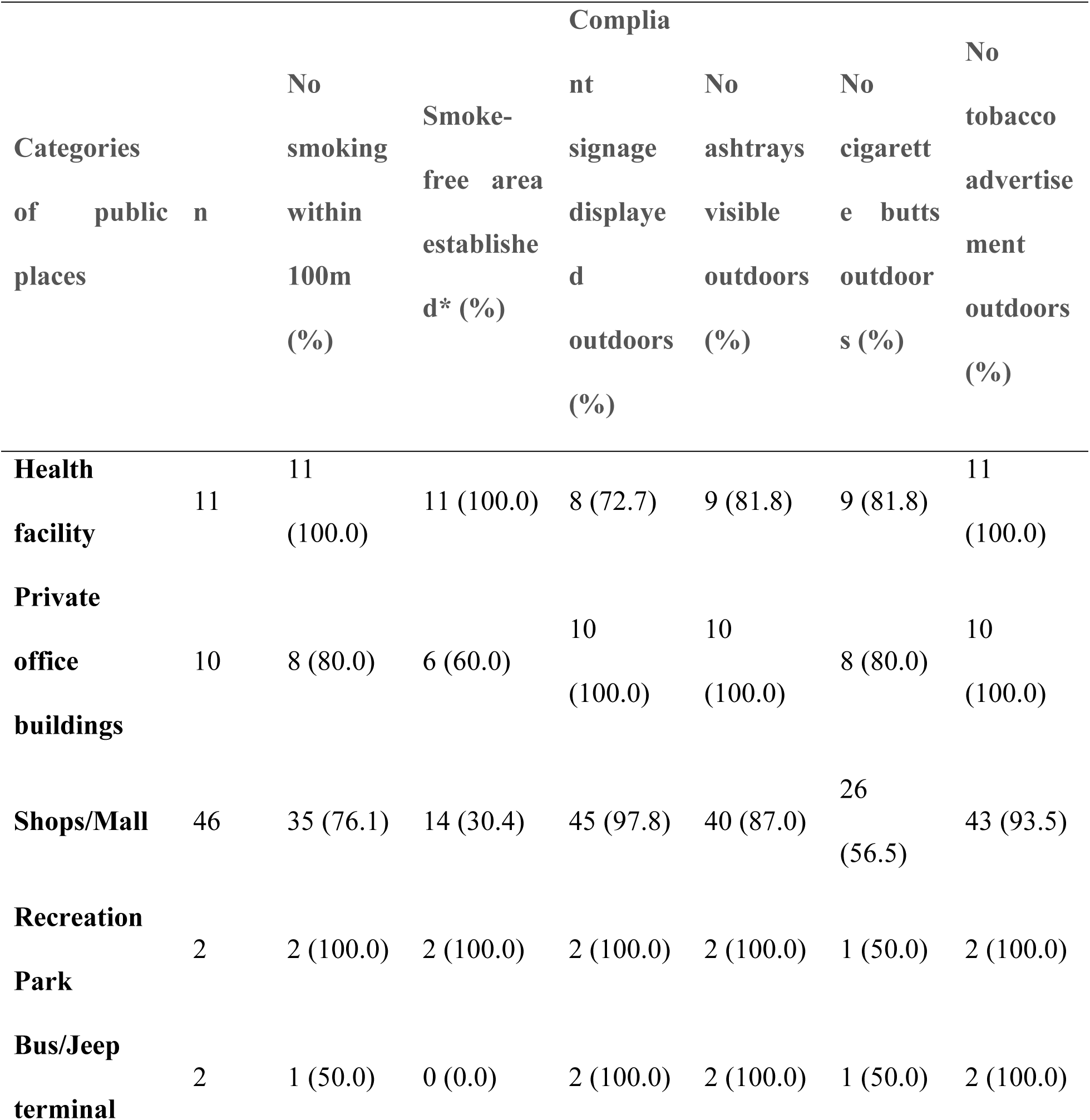
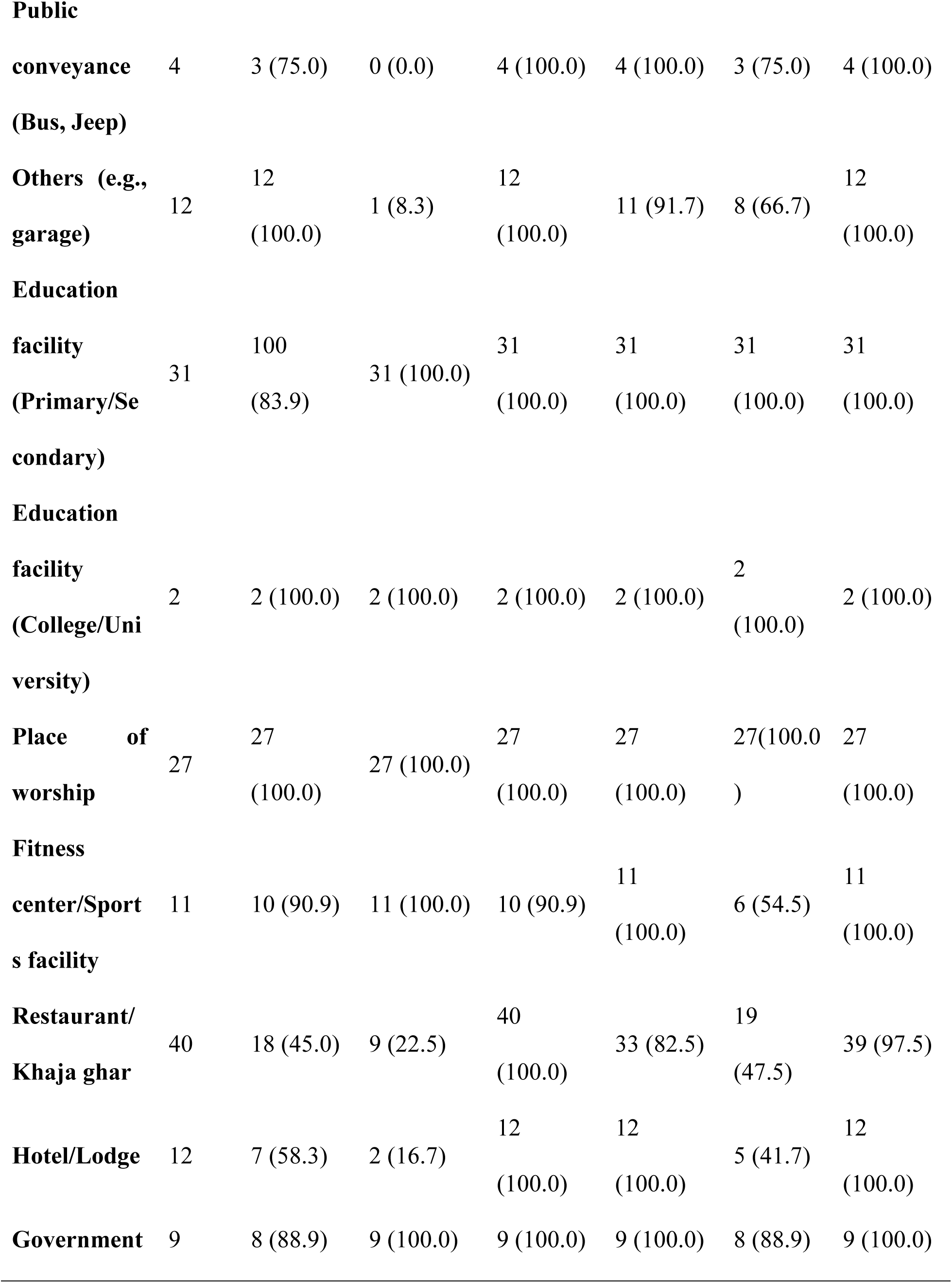
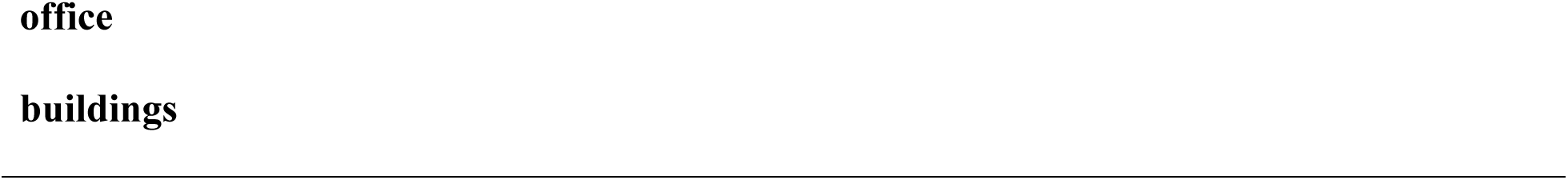
Compliance with smoke-free laws for outdoor observations across different public places.

### Factors Associated with Smoking Behavior in Public Places

According to table 4, the presence of visible ashtrays showed strong, statistically significant associations with smoking behavior. Venues with visible indoor ashtrays had 16 times higher odds of smoking being observed (OR=16.10, p<0.001), while those with outdoor ashtrays had 7.5 times higher odds (OR=7.48, p<0.001). In contrast, ‘No Smoking’ signage showed no statistically significant associations, though venues with indoor signage demonstrated perfect compliance (0% smoking prevalence).

**Table 4:**
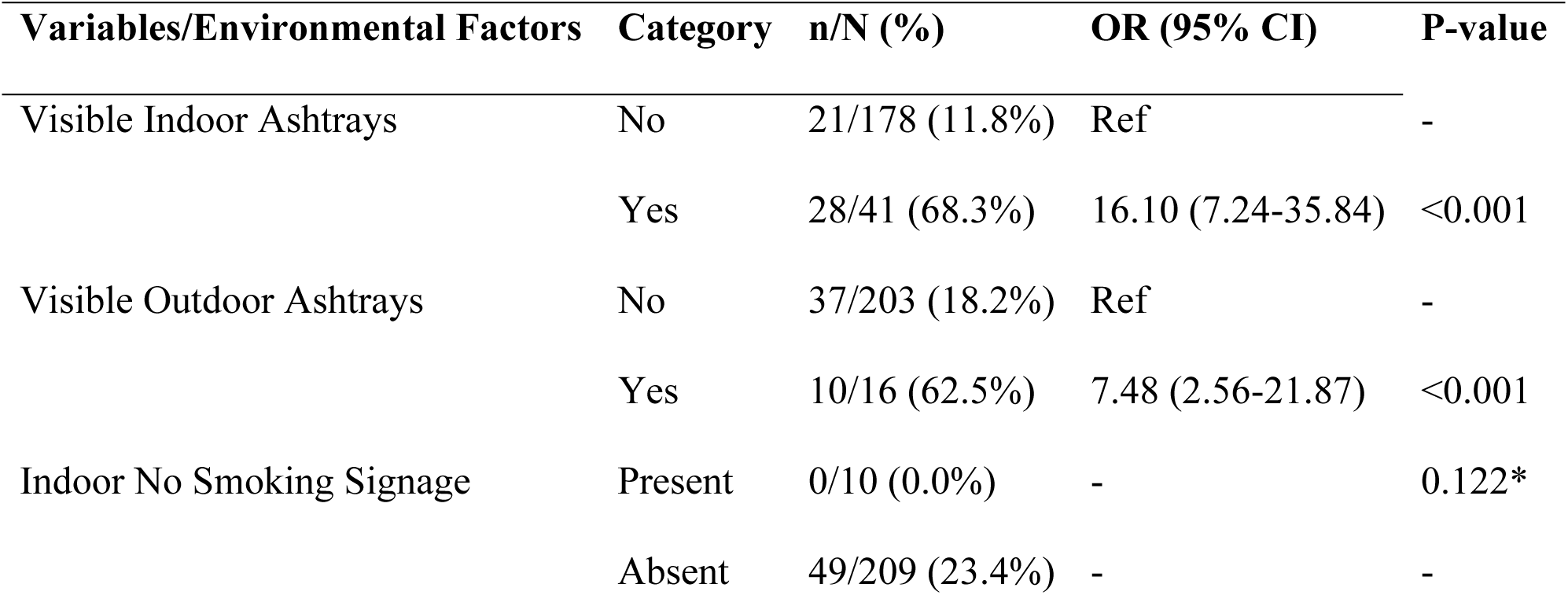
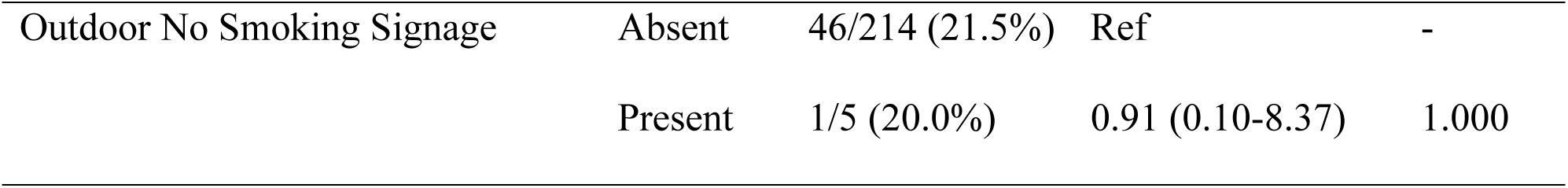
Factors Associated with Smoking Behavior in Public Places (N=219)

## DISCUSSION

This study assessed compliance with smoke-free legislation in Purbakhola Rural Municipality, Nepal, and found mixed results. Compliance was high in formal institutional settings such as health facilities, educational institutions, and government offices, whereas hospitality sectors (e.g. restaurants and hotels) showed lower compliance. In the hospitality sectors, the study demonstrated indoor smoking, the presence of ashtrays, and cigarette butt litter. The lack of no-smoking signage was the main gap, in almost all types of public places. These findings highlight a critical implementation gap between legislation and practices in local levels particularly rural Nepal.

Our results are consistent with international and regional studies showing that compliance tends to be highest in formal institutional settings and lowest in hospitality sectors. The studies in Turkey and United States of America found that hospitals and government buildings showed near-universal compliance, but restaurants and bars consistently lagged behind, with frequent indoor smoking and a lack of signage (14–16). In Russia, higher rates of non-compliance were also documented in restaurants, cafés, bars, and nightclubs, as well as in common areas of apartment buildings and indoor workplaces. By contrast, violations were less frequently observed on public transport, in government buildings, health and sports facilities, and educational institutions such as colleges and universities in Russia(17). Similarly, observational research in Karachi reported that only 47% of restaurants were free of active smoking and 39% displayed appropriate signage, figures strikingly similar to our study’s findings of 45% and 37.5%, respectively(18). In Punjab, India, observed active smoking was in 34% of public places despite legal prohibitions, with restaurants being the most non-compliant category(19). The Bangladesh study also reported relatively low overall compliance, with hospitality venues accounting for the majority of violations(20). In a previous study conducted in urban areas of Nepal, the highest compliance (75.0%) was observed in government office buildings, while the lowest compliance (26.3%) was reported in eateries, entertainment, and shopping venues (21–23). These parallels reinforce that hospitality settings are consistently the weakest point in smoke-free law enforcement across South Asia(24).

The presence of visible ashtrays was the strongest predictor of smoking behavior. Venues with indoor ashtrays had over sixteen times higher odds of observed smoking, and those with outdoor ashtrays had nearly 7.5 times higher. This indicates that ashtrays are not merely containers for tobacco ashes but powerful environmental cues that normalize and facilitate smoking, directly contributing to the high non-compliance observed in hospitality venues. Conversely, the role of ‘No Smoking’ signage remains complex. Our analysis found no statistically significant protective effect for signage. This may be attributed to the small sample size, as only 10 out of 219 indoor public places had no-smoking signage. However, a promising signal was noted: not a single instance of smoking was observed in the few venues that had indoor signage displayed. This suggests that where signage is implemented, it may be effective.

The absence of signage in our study is particularly concerning, as signage plays a vital role in raising awareness, signaling legal restrictions, and empowering public self-enforcement(21). The widespread absence of compliant signage, observed in over half of health facilities and all government offices, aligns with evidence from Bangladesh, Guatemala, and Ethiopia. This suggests that signage non-compliance is a recurrent systemic failure, even in settings with high overall institutional compliance(25, 26). The lack of signage may undermine smoke-free policies by fostering low public awareness, despite overwhelming support for such measures. This demonstrates that signage is a crucial operational tool for compliance, not merely a symbolic one. Another recurring theme across multiple studies is the role of cultural norms and enforcement. In Nepal, as in India and Bangladesh, smoking in restaurants and hotels appears to be normalized, with weak enforcement mechanisms enabling continued non-compliance and both emphasized that laws alone are insufficient; enforcement, monitoring, and penalties are critical to changing behavior(19, 20). The persistence of ashtrays and cigarette butt litter in hospitality venues indicates not only a lack of active enforcement but also tacit acceptance of smoking by business owners. This finding aligns with evidence from Karachi, where restaurant owners expressed fear of losing customers if smoking was strictly prohibited (18).’

Our findings therefore have clear policy implications. First, mandating the signage across all public venues utmost importance and it is actually a simple, low-cost intervention with high potential impact. Previous research has shown the presence of signage works as the means of that awareness of tobacco-free policies within institutions((27). Second, enforcement must extend beyond government institutional venues to hospitality settings, where second-hand smoke exposure is greatest. Finally, local governments should integrate tobacco control into routine monitoring and enforcement structures, supported by public awareness campaigns that target low-knowledge groups such as small shopkeepers and hospitality business owners and managers.

### Strengths and Limitations

This study is among the first to assess compliance with smoke-free laws in a rural municipality of Nepal, providing granular evidence across 14 categories of public places. The use of direct observation and photographic evidence strengthens the reliability of findings. However, the study was limited to one municipality and may not be generalizable to all rural areas of Nepal. Additionally, data collection during daytime business hours may underestimate non-compliance in restaurants and hotels, where evening smoking could be more prevalent.

## CONCLUSIONS

This study demonstrates a significant implementation gap in smoke-free law compliance within Purbakhola Rural Municipality. While institutional settings like health facilities and schools showed near-perfect adherence, the hospitality sector exhibited major deficits, with high rates of active indoor smoking in restaurants (55.0%) and hotels (41.7%). The most systemic failure was the lack of compliant no-smoking signage, which was absent in over half of health facilities and all government offices. Given the near-universal public support for these laws, these compliance gaps are not due to public opposition but to inadequate enforcement and awareness.

To bridge this policy-practice divide, a targeted, multi-strategy approach is essential. We recommend Purbakhola Rural Municipality immediately prioritize the universal installation of standardized signage across all public places, led by government example. Concurrently, enforcement must be strengthened through the formation of a multi-sectoral monitoring committee to conduct unannounced inspections and impose consistent penalties, with a specific focus on hospitality venues. Furthermore, targeted awareness campaigns via local media should be launched to educate business owners in low-compliance sectors and the general public. For long-term impact, tobacco control must be mainstreamed into the municipality’s annual health plans with a dedicated budget. Finally, point-of-sale regulations near schools must be strictly enforced to prevent illegal advertising and sales. By implementing these measures—focusing on signage, enforcement, awareness, and integration—the municipality can transform its strong legal framework into effective public health protection for its residents.

## Data Availability

The data is uploaded as Supporting information.

## Acknowledgement

We acknowledge office of Purbakhola local municipality for its support. We also used ChapGpt to correct the English language only.

## Ethical approval

Not application because no human studies were applied. **Supporting Information**: The data is uploaded as supporting information.

## Author Contributions

Conceptualization: Bhakta Bahadur KC, Ananda Bahadur Chand Curation: Bhakta Bahadur KC, Amit Bam

Formal Analysis: Bhakta Bahadur KC, Tara Singh Bam Investigation: Amit Bam, Ananda Bahadur Chand Methodology: Bhakta Bahadur KC, Tara Singh Bam Project administration: Ananda Bahadur Chand, Amit Bam

Resources: Ananda Bahadur Chanda, Tara Singh Bam

Software: Bhakta Bahadur KC, Amit Bam Supervision: Ananda Bahadur Chand, Amit Bam

Validation: Bhakta Bahadur KC, Ananda Bahadur Chand. Tara Singh Bam Visualization: Bhakta Bahadur KC, Amit Bam, Tara Singh Bam.

Writing – original draft: Bhakta Bahadur KC

Writing – review & editing: Bhakta Bahadur KC, Ananda Bahadur Chand, Amit Bam, Tara Singh

